# Global performance of Human Papillomavirus Typing, Screening, and Evaluation as assessed using Proficiency Panels traceable to International Standards

**DOI:** 10.64898/2026.01.03.26343391

**Authors:** Laila Sara Arroyo Mühr, Emel Yilmaz, Camilla Lagheden, Carina Eklund, Joakim Dillner

## Abstract

**Background:** Accurate and comparable human papillomavirus (HPV) testing is essential for HPV vaccine research, HPV surveillance, and cervical cancer screening. In 2008, the WHO HPV Laboratory Network initiated global HPV proficiency testing traceable to International Standards (IS), which has been issued regularly since. Here, we summarize recent results and trends over time.

**Methods:** The HPV genotyping panel, used since 2008, consists of 43 blinded samples, including HPV 6, HPV 11, all oncogenic and vaccine-targeted HPV types, as well as extraction controls. A smaller screening panel, launched in 2022, includes 13 blinded samples designed for cervical screening needs. Laboratories worldwide test the panels using their own methods, and results are centrally evaluated at the International HPV Reference Center. Proficiency has hitherto been defined as absence of false positives, detection of HPV16/18 at 10 IU/µL, and of other oncogenic types at 100 IU/µL (genotyping) or 1000 IU/µL (screening).

**Results:** Participation in the genotyping panel increased from 54 laboratories in 2008 to >130 in 2021. Proficiency rose from ∼25% in early years to >80% by 2024, with >99% correct detection for most genotypes. Optional low-copy challenges (e.g., 1 IU/µL HPV16/18) were detected by >95% of laboratories by 2024. Screening panel participation increased from 84 laboratories in 2022 to 132 in 2024, with overall proficiency improving from 77% to 95% and >96% datasets free of false positives.

**Conclusions:** The global HPV proficiency program enables global laboratory quality assurance. Annual proficiency testing is essential for high-performance HPV testing and evaluation supporting cervical cancer elimination.

**summary:** Global HPV proficiency program traceable to international standards shows marked improvement from 2008–2024. Participation expanded worldwide, false positives decreased, and most HPV types reached near-universal detection. Annual proficiency testing supports HPV research, surveillance, screening and evaluation of novel assays.

## Background

Accurate and internationally comparable human papillomavirus (HPV) DNA testing is fundamental for both scientific and public health purposes.(1) Reliable HPV genotyping underpins vaccine research, studies of viral epidemiology, and monitoring of vaccination program impact, while sensitive and specific detection of oncogenic HPV types forms the basis of cervical cancer screening worldwide.(2–4) Standardized quality assurance is therefore essential to ensure that the rapid expansion of HPV testing technologies yields internationally comparable results, across laboratories, countries, and over time.(1, 2)

The World Health Organization (WHO) established the Global HPV Laboratory Network (HPV LabNet) in 2005.(1) The goal of the HPV LabNet was to harmonize and standardize laboratory procedures used in HPV vaccinology including building internationally comparable quality assurance systems, development of international standards (IS) and reference reagents, laboratory manuals, and establishment of external quality assessment programs.(1)

The global HPV proficiency panels are blinded sets of challenge samples containing none, one, or several HPV plasmids in a background of human DNA. The panels are designed by the International HPV Reference Center (IHRC) and distributed to laboratories worldwide. (5) Any laboratory can participate using its own HPV testing methods, and results are centrally evaluated, providing a shared benchmark for sensitivity (detecting HPV when present) and specificity (avoiding false positives).(6)

The HPV genotyping proficiency panels were designed to evaluate full genotyping performance, which is crucial for HPV vaccinology and surveillance. The HPV screening proficiency panels focus on the needs of cervical cancer screening, where high sensitivity for the most oncogenic HPV types (16/18) and strict avoidance of false positives is paramount.(7)

Technical reports from 2008–2024 are published on the IHRC website (https://www.hpvcenter.se/proficiency_panel/) and, for 2008–2023 have been summarized in the scientific literature(5–13) except the genotyping study in 2017 and the screening study in 2023 that are first reported here. Initial criteria for the screening panel essentially mirrored those of the genotyping panel, but studies on virus amounts in screening samples taken before cervical cancer (14) have spurred ongoing international collaborative studies aiming to better define which analytical limit of detection thresholds that are medically meaningful

Together, the screening and genotyping panels provide a comprehensive framework for global quality assurance of HPV testing, supporting research, surveillance, screening, and analytical evaluation of new HPV testing technologies.

## Methods

### The HPV Genotyping proficiency panel

The genotyping proficiency panel was the first external quality assurance tool established by the WHO HPV LabNet, designed to provide laboratories with a standardized and reproducible means of evaluating their ability to correctly identify individual HPV types with results traceable to international standards. The first panel was launched in 2008, following the establishment of WHO International Standards for HPV16 and HPV18 DNA.(5) Since then, each panel has consisted of 43 blinded samples, including one negative control, 34 single-type samples at varying concentrations, and eight multi-type mixtures. Three cell line suspensions are also included as extraction controls, consisting of dilution series of HPV16-positive SiHa cells and HPV-negative C33A cells.(5, 6, 8–12, 15)

The panel includes all oncogenic and probably oncogenic HPV types (HPV16, 18, 31, 33, 35, 39, 45, 51, 52, 56, 58, 59, and 68a/68b) and the non-oncogenic vaccine types (HPV6, 11). All plasmids contained full-length viral sequences except for the plasmid HPV68a, which consisted only of the L1 gene. The initial panel distributed in 2005 revealed that many assays failed to detect HPV68a, likely because their primers and probes had been designed against the HPV68b sequence. Consequently, HPV68b plasmids were added from 2010 onwards to address this issue. HPV66, originally considered oncogenic, but later reclassified by IARC as non-oncogenic, was initially included in the proficiency panels, but removed after 2018 when new evidence had emerged and HPV66 was classified as not oncogenic.(16)

Laboratories worldwide were invited to participate using their own testing methods. Results were decoded and evaluated by the IHRC. Proficiency criteria required correct identification of HPV16 and HPV18 at 10 IU/µl and of other types at 100 genome equivalents (GE)/µl in accordance with the consensus requirements established at a WHO workshop in 2009.(17) Since 2019, 100% specificity (no false positives) has been required, and the same criterion is here applied retrospectively to older data. In addition to the concentrations required for proficiency, samples containing low-copy challenges (10 or 1 GE) and multi-type mixtures were included to provide further assessment of assay performance, although correct detection was not required for proficiency. The detailed composition of each genotyping proficiency panel, including plasmid types, concentrations, and control materials, is described in a series of peer-reviewed articles and in technical reports prepared by the IHRC.(5–12) For every panel distributed, the IHRC issues a technical report summarizing results in detail, with summaries published in scientific literature. All technical reports are publicly available at https://www.hpvcenter.se/proficiency_panel/.

### The HPV Screening proficiency panel

The screening proficiency panel was introduced in 2022 to meet the specific requirements of HPV testing in cervical cancer screening, which differ from those of HPV testing in research and vaccinology. Whereas the typing panel was designed to assess full genotyping performance, the screening panel focused on ensuring very high sensitivity for HPV16 and HPV18, reliable detection of other oncogenic types, and strict avoidance of false positives. To reduce workload for laboratories, the screening panel was designed as a compact set of twelve blinded samples in its first year and expanded to thirteen samples in subsequent rounds.

The first panel was distributed in 2022 and included samples containing HPV16, HPV18, HPV31, HPV33, HPV45, HPV52, HPV58 and a pooled sample of other oncogenic types (HPV35, 39, 51, 56, 59, 68), in addition to a negative control.(7, 13) Proficiency was defined as detection of at least 10 IU/ul of HPV16/18, 1000 IU/ul of HPV31, 33, 45, 52, and 58, and the absence of false positives, mirroring the criteria of the typing panel. Mixed-genotype pools were included as optional challenges containing 100 GE/ul each, as were low-concentration samples containing 1 IU/ul of HPV16/18.

The 10 IU/µL concentration of HPV16 and HPV18 was not detected by any laboratory that used the Hybrid Capture 2 (HC2) assay, which has a manufacturer-stated detection limit of approximately 100 copies/µL. (13). In 2023, a 100-copy/µL (1.08 pg/mL as specified by the manufacturer) sample for HPV16 and HPV18 was therefore added as a service to laboratories using HC2.(18) However, as the number of laboratories using HC2 rapidly decreased, these 100-copy samples were removed from later panels. In response to the WHO Target Product Profile (TPP), (19) which requires detection of eight major oncogenic types, namely HPV16, 18, 31, 33, 35, 45, 52, and 58, the 2024 version of the panel included HPV35 as an individual sample due to its high prevalence in cervical cancers from sub-Saharan Africa.(20)

As for the genotyping panels, results from the screening panels are decoded and evaluated at the IHRC. The detailed composition of each panel, together with technical reports summarizing performance, are available at https://www.hpvcenter.se/proficiency_panel/. The findings from the first screening panel were summarized and published as a peer-reviewed article.(7)

## Results

### The Genotyping proficiency Panel

#### Participation

Participation in the typing panel varied over time (Figure 1). Following a steady rise between 2008 and 2014, when the number of participating laboratories increased from 54 to 121, the program maintained high engagement through 2017. A sharp decline occurred in 2019, when only 78 laboratories submitted 110 datasets, coinciding with a drop in overall proficiency. This concerning finding prompted a major investment from the Gates Foundation to support an enhanced, annually issued program. As a result, participation rebounded strongly in 2021, reaching 132 laboratories and 211 datasets - the highest level recorded to date. In subsequent years, participation stabilized between 78 and 96 laboratories, reflecting the concurrent introduction of the HPV screening panel in 2022. Laboratories focused on epidemiology and vaccinology continued with the genotyping panel, whereas those engaged in cervical screening predominantly transitioned to the screening panel.

**Figure 1:**
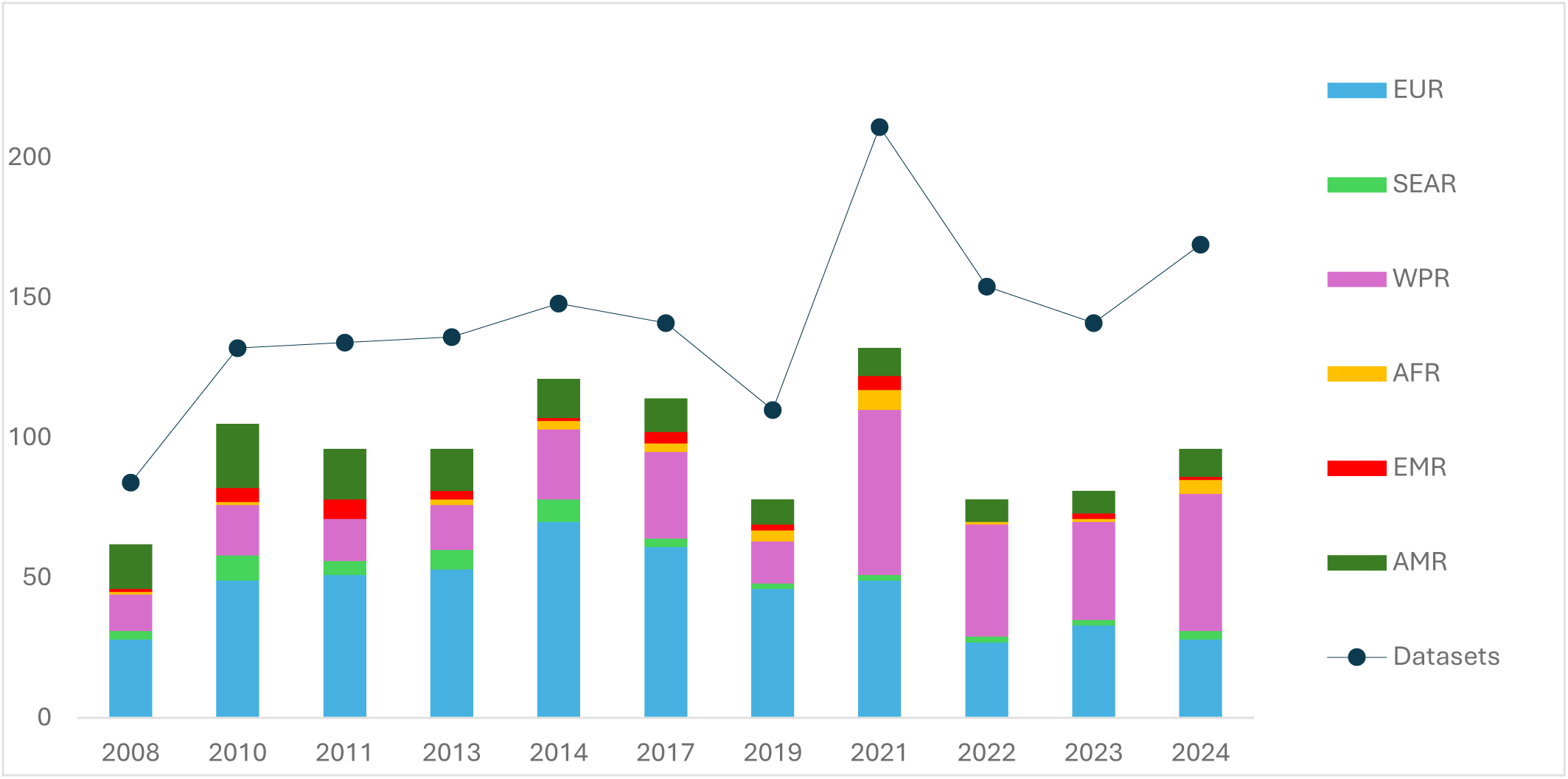
Participation in HPV genotyping proficiency panels, 2008–2024. Global participation in the HPV typing proficiency studies. Bars show the regional distribution of participating laboratories by WHO region (EUR: Europe; SEAR: South-East Asia; WPR: Western Pacific; AFR: Africa; EMR: Eastern Mediterranean; AMR: Americas). The line indicates the total number of datasets submitted each year.

#### Required concentrations – proficiency by HPV type

Proficiency by HPV type has steadily improved over time and now approaches 100% correct detection for nearly all types. For the most carcinogenic types, HPV16 and HPV18, performance was already high in the earliest panels, with 94.9% and 92.2% of datasets correctly identifying them in 2008. These rates continued to increase, reaching 99.4% and 100%, respectively, by 2024 (Figure 2A).

**Figure 2:**
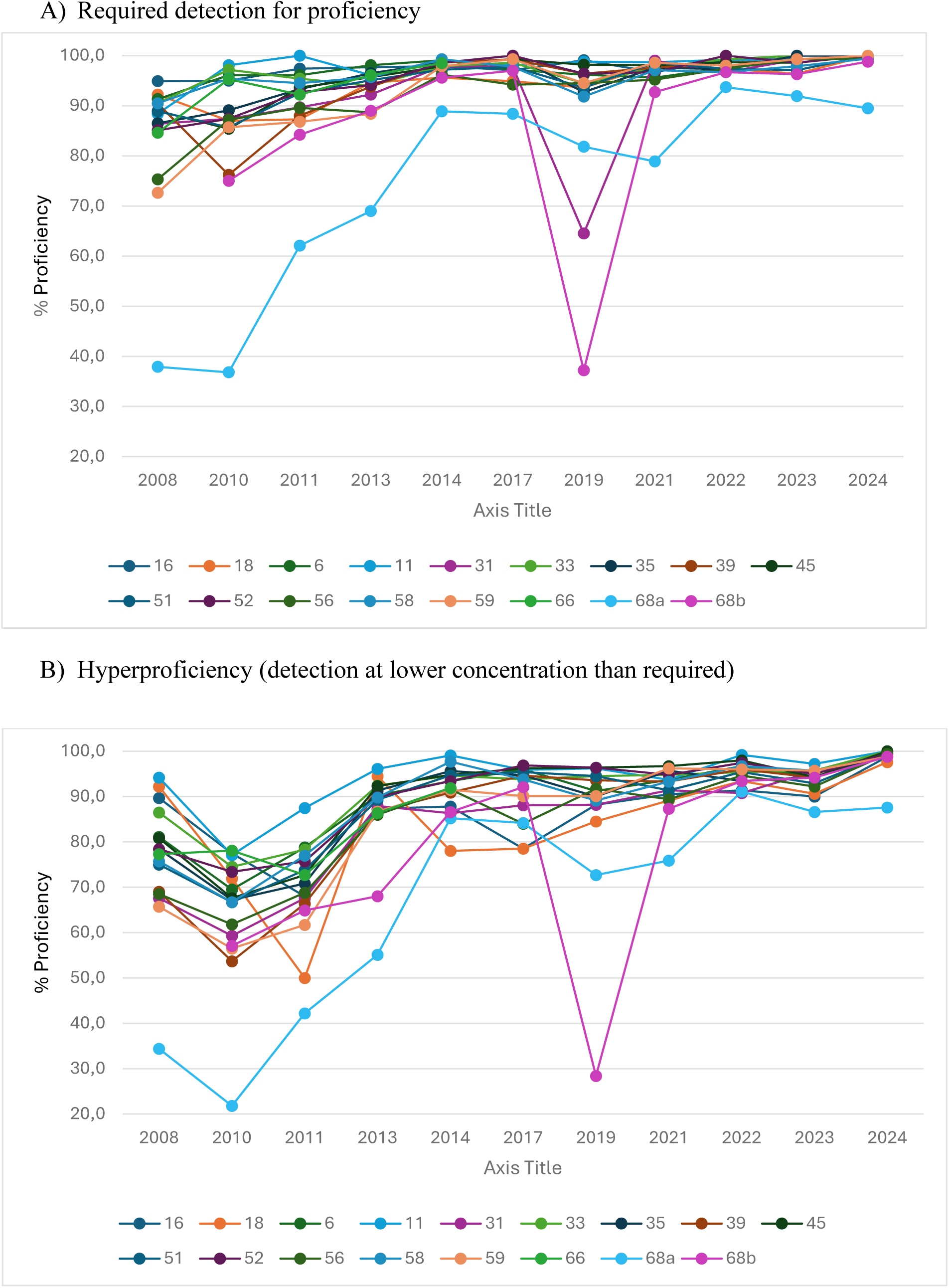
Percentage of datasets reporting correct HPV type as claimed and hyper with no false positive HPV type detected.

For other oncogenic types, proficiency was lower in the early years. HPV31, for instance, was correctly detected in 86.4% of datasets in 2008 even declining to 64.5% in 2019 but reached 100% by 2024. Similar trends were observed for HPV33 (90.5% in 2008, >99% from 2022 onward), HPV45 (89.0% in 2008, 100% in 2024), and HPV52 (85.1% in 2008, >96% from 2014 onward). The non-oncogenic vaccine types HPV6 and HPV11 also showed progressive improvement, achieving 100% proficiency by 2024 (Figure 2A).

HPV56 and HPV59 presented greater difficulty initially, with correct detection in only 75.3% and 72.6% of datasets in 2008, respectively. By 2014, both exceeded 96% proficiency and reached 99.4% and 100% by 2024.

The most persistent challenge was HPV68a. In 2008, only 37.9% of datasets detected it. Although performance improved over time, it remained below that of other types, reaching 89.5% by 2024. In contrast, HPV68b (introduced in 2010) performed well from the outset (75.0% in 2010, >84% in 2011) and exceeded 95% between 2013 and 2017. A sharp decline occurred in 2019 (37.2%), but proficiency recovered thereafter (92.7% in 2021; 98.8% in 2024). HPV66, included until 2017, consistently performed well (84.6% in 2008; 98.5% in 2014).

By 2024, proficiency across all HPV types at required concentrations exceeded 99%, except for HPV68a (89.5%) and HPV68b (98.8%), demonstrating substantial strengthening of global laboratory capacity for reliable HPV DNA testing across genotypes. (Figure 2A)

#### Optional detection of low concentration HPV – “hyper proficiency”

Detection of low-concentration samples, where positivity was not required for proficiency, improved markedly over time. In the earliest panels (2008–2011), “hyper proficiency”, defined as detection of HPV below the required threshold, was generally low (<80% for most types). For example, HPV18 at 1 IU/µL was detected in only 71.9% of datasets in 2010 and 50.0% in 2011 whereas HPV31, HPV59, and HPV68a, tested at 10 IU/µL, were detected in 59.3%, 56.5%, and 21.8% of datasets, respectively. HPV68b, introduced in 2010, also showed limited early performance, with only 57.1% hyper proficiency (Figure 2B).

By 2013–2014, performance had improved substantially, with most genotypes exceeding 85% detection except for HPV18, HPV68a, and HPV68b (78% for HPV18 in 2014). From 2014 onward, detection at low concentrations continued to strengthen. By 2022, all types exceeded 90% hyper proficiency, with additional gains in 2023 and 2024. In the most recent round (2024), nearly all HPV types reached or approached 100% detection at optional concentrations, with only HPV68a (87.6%) remaining below full proficiency. HPV68b, which had fluctuated in earlier years, recovered strongly and achieved 98.8% in 2024 (Figure 2B).

#### Overall proficiency, false positivity and type of assays

The first panels in 2008–2010 showed that only about one-quarter of datasets were fully proficient, and approximately half of laboratories (42.5% and 50.3% in 2008 and 2010, respectively) reported at least one false positive, underscoring the importance of external quality assessment. From 2011 to 2017, performance improved, with 40.3–68.1% of datasets being fully proficient and false positivity decreasing to 30% of datasets (Table 1).

**Table 1:**
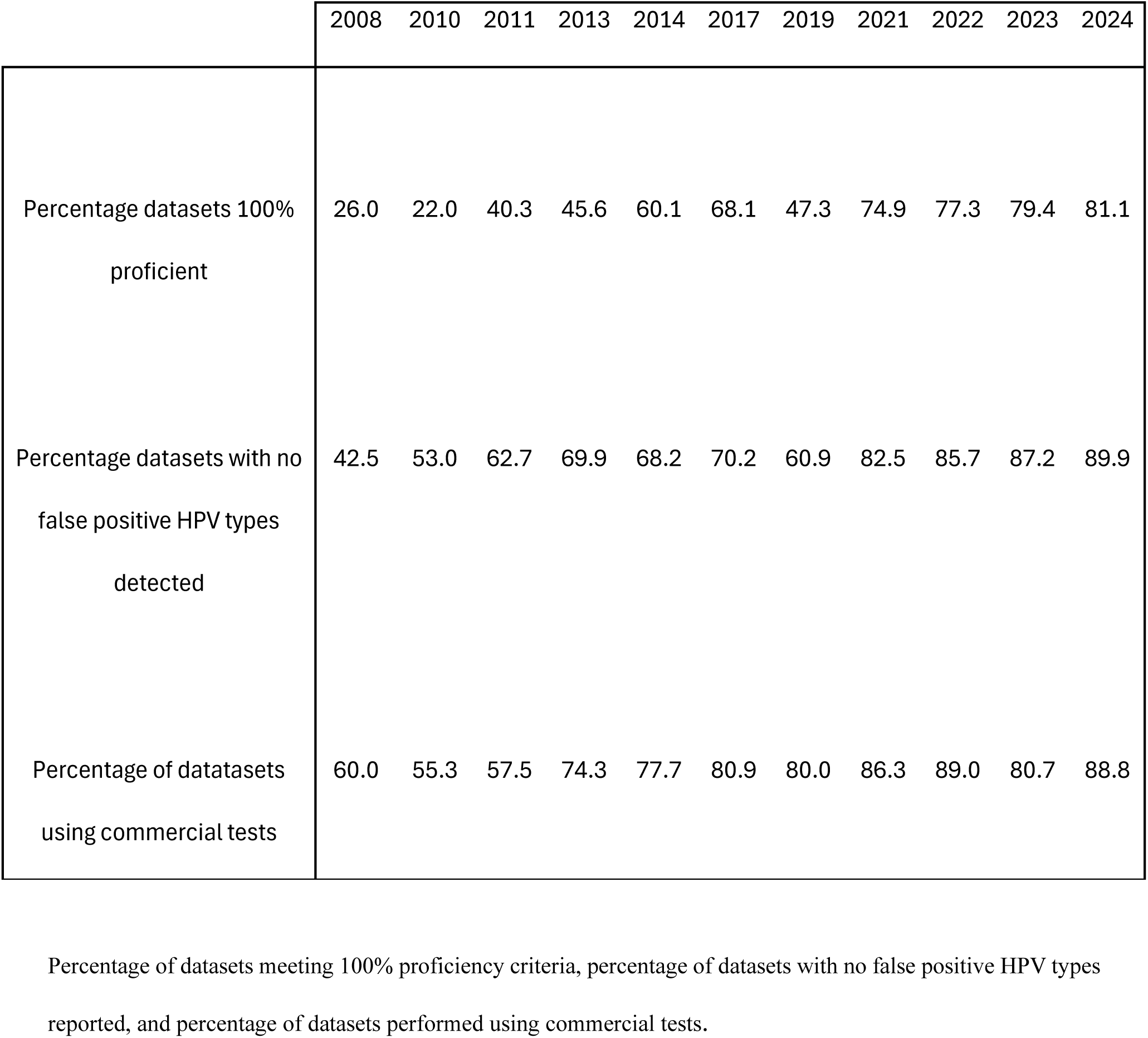
Trends in laboratory performance and assay use in the HPV genotyping proficiency panels, 2008–2024.

The 2019 panel provided an unexpected wake-up call: after years of improvement, the overall proficiency declined sharply, with only 42% of datasets being fully proficient and one-quarter reporting false positives. This highlighted an urgent need for improved quality assurance. Beginning in 2021, after annual distribution of panels was reintroduced, performance improved again. The 2021 panel recorded the highest proficiency since the program began, with 75% of datasets fully proficient and more than 82% reporting no false positives. This upward trend continued, with 81% fully proficient datasets and almost 90% false-positive–free datasets in 2024 (Table 1).

The use of commercial assays has increased steadily, reaching nearly 89% of datasets in 2024, indicating widespread adoption of standardized testing platforms.

#### Laboratories that participated in 2024 and in the years 2008, 2010, 2011, 2013, 2014, 2017, 2019, 2021, 2022 and 2023

There were 62 laboratories that participated in the last proficiency panel (2024) that had also participated in the HPV LabNet proficiency panels from at least one previous year. Ten laboratories that submitted results in 2024 had participated already in 2008 at the start of the proficiency studies. While some of the laboratories used the same tests during all years, other laboratories have changed at least one of the tests used. Proficiency increase is seen in Table 2.

**Table 2:**
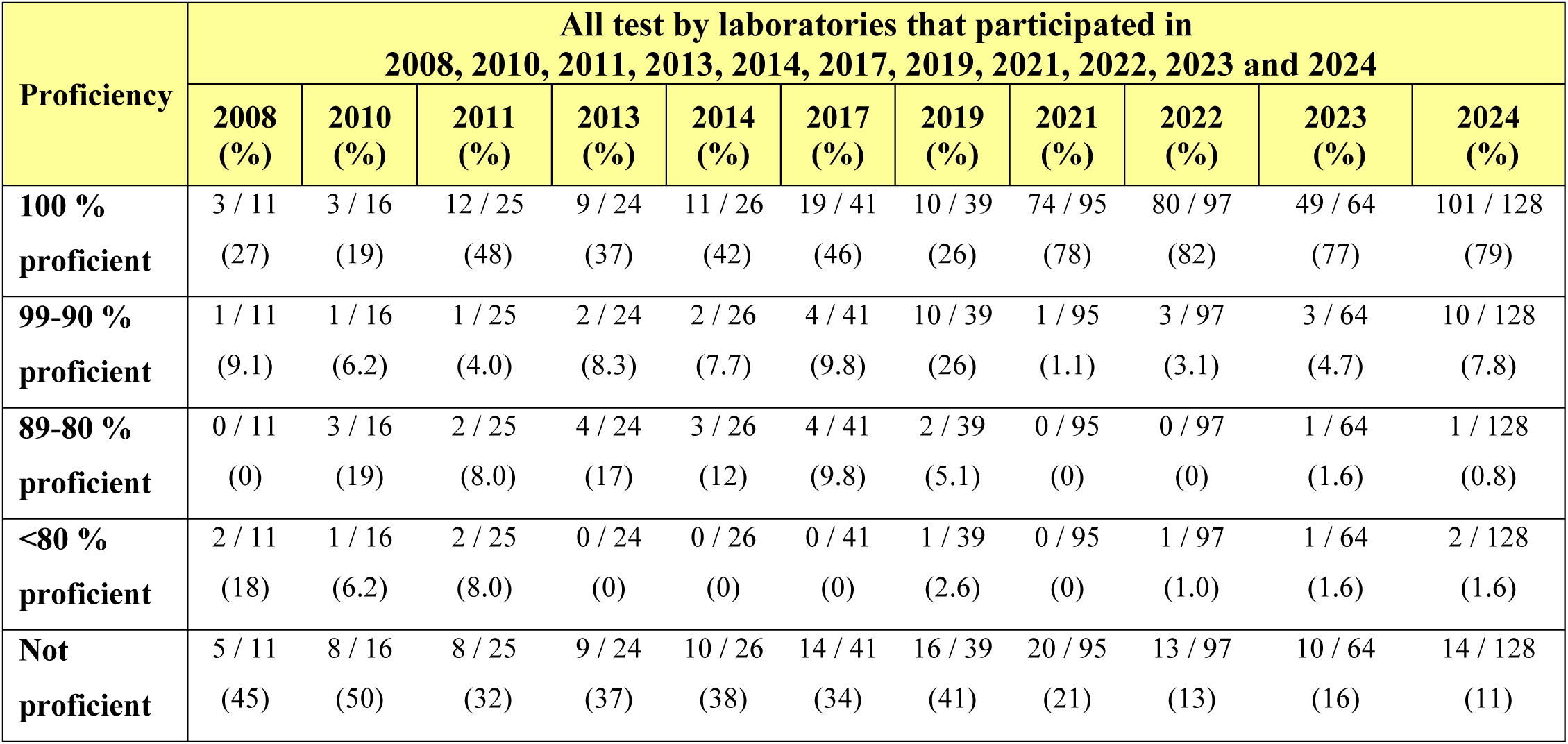
Proficiency of detecting HPV types by laboratories that participated in 2024 PP, with data from 2008, 2010, 2011, 2013, 2014, 2017, 2019, 2021, 2022 and 2023.

### The Screening proficiency panel

The first screening proficiency panel was distributed in 2022. Participation expanded rapidly, with 84 laboratories submitting 158 datasets in 2022, 95 laboratories and 152 datasets in 2023, and 132 laboratories and 208 datasets in 2024 (Table 3). Geographic coverage also broadened, with increasing participation from Africa and the Eastern Mediterranean. Most datasets were generated using commercial assays, which rose from 80% in 2022 to more than 97% in 2024.

**Table 3:** Participation, assay use, and proficiency in the HPV screening proficiency panels, 2022–2024.

| Screening proficiency studies | 2022 |  | 2023 |  | 2024 |  |
| --- | --- | --- | --- | --- | --- | --- |
| Participation | Laboratories | Datasets | Laboratories | Datasets | Laboratories | Datasets |
| Total | 84 | 158 | 95 | 152 | 132 | 208 |
| PAHO | 27 | 35 | 32 | 34 | 49 | 53 |
| EURO | 29 | 36 | 45 | 55 | 52 | 63 |
| AFRO | 0 | 0 | 1 | 1 | 4 | 4 |
| EMRO | 0 | 0 | 0 | 0 | 2 | 2 |
| SEARO | 2 | 2 | 0 | 0 | 1 | 1 |
| WPRO | 26 | 85 | 17 | 62 | 24 | 85 |
| Type of assays used in datasets |  |  |  |  |  |  |
| Commercial | 133 (84.2%) |  | 145 (95.4%) |  | 202 (97.1%) |  |
| In house | 25 (15.8%) |  | 7 (4.6%) |  | 6 (2.9%) |  |
| Overall Proficiency of datasets |  |  |  |  |  |  |
| 100% Proficient | 128/158 (81.0%) |  | 138 (90.8%) |  | 197 (94.7%) |  |
| No false positivity | 150/158 (94.9%) |  | 146 (96.1%) |  | 200 (96.2%) |  |
| Proficiency of each sample |  |  |  |  |  |  |
| HPV 16 1 IU/ul | 99.4 | 157/158 | 95.4 | 144/151 | 95.2 | 197/207 |
| 10 IU/ul | 86.1 | 136/158 | 97.4 | 147/151 | 99.0 | 205/207 |
| 100 IU/ul | - | - | 98.7 | 149/151 | - | - |
| HPV18 1 IU/ul | 99.4 | 157/158 | 82.7 | 124/150 | 96.6 | 201/208 |
| 10 IU/ul | 86.1 | 136/158 | 96.7 | 145/150 | 99.0 | 205/207 |
| 100 IU/ul | - | - | 98.7 | 148/150 | - | - |
| HPV31 1000 GE/ul | 98.1 | 155/158 | 99.3 | 149/150 | 100 | 208/208 |
| HPV33 1000 GE/ul | 98.1 | 155/158 | 99.3 | 150/151 | 99.5 | 206/207 |
| HPV35 1000 GE/ul | - | - | - | - | 99.5 | 207/208 |
| HPV45 1000 GE/ul | 93.0 | 147/158 | 99.3 | 151/152 | 100 | 206/206 |
| HPV52 1000 GE/ul | 96.2 | 152/158 | 100 | 149/149 | 100 | 207/207 |
| HPV58 1000 GE/ul | 97.5 | 154/158 | 99.3 | 149/150 | 100 | 206/206 |
| 31/33/45/52/58* 100 GE/ul | 98.7 | 156/158 | 97.3 | 145/149 | 99.0 | 204/206 |
| 35/39/51/56/59/68* 100 GE/ul | 98.1 | 155/158 | 96.7 | 145/150 | 98.6 | 205/208 |
| Negative 0 | 99.4 | 157/158 | 99.3 | 151/152 | 99.5 | 207/208 |

Overall proficiency improved markedly across rounds. In 2022, 81.0% of datasets met all proficiency criteria and 94.9% reported no false positives. By 2023, proficiency had increased to 90.8%, and by 2024 reached 94.7%, with 96.2% of datasets showing no false positives.

Performance by HPV type followed similar trends. At the required concentrations, detection of HPV16 and HPV18 rose from approximately 86% in 2022 to more than 99% in 2024. The lower performance in the first round was largely attributable to the HC2 assay, which was unable to detect HPV16/18 at the required threshold of 10 IU/µL, consistent with the manufacturer’s stated detection limit of 100 copies. (18) To accommodate this limitation, a 100-copy HPV16/18 sample was introduced in 2023, allowing HC2 to perform within its specifications. However, as HC2 was subsequently phased out by most laboratories, this adjustment was discontinued in the 2024 panel. For other oncogenic types, proficiency already exceeded 93% in the first round and reached 100% for most by 2024. HPV35 was included as an individual sample in 2024 to comply with new requirements from the WHO Target Product Profile and was detected with >99% proficiency.(19) Mixed-genotype pools and negative controls also performed consistently well, with >99% correct results in the most recent panels.

## Conclusions

Over more than fifteen years, the global HPV proficiency program has provided unique insights into the state of HPV testing worldwide. Participation has expanded steadily, underscoring the value that laboratories place on external quality assessment and their commitment to producing internationally comparable results. The panels have become not only a reference point for quality assurance but also a means of tracking the evolution of HPV testing methods and performance over time.

The results of the program demonstrate that proficiency cannot be taken for granted. The decline in performance observed in 2019, when nearly one-third of laboratories reported at least one false positive result, highlighted how rapidly quality can deteriorate if not continuously monitored.(12) The sharp recovery documented in 2021, after annual distribution of panels resumed, emphasized the importance of regular participation as a safeguard for laboratory quality.(8) By enabling early detection and correction of problems, annual testing has proven essential for maintaining robust global performance.(6)

Another consistent lesson has been that both sensitivity and specificity are critical.(2, 21) While high analytical sensitivity for oncogenic HPV types is indispensable, avoidance of false positives is equally important, particularly in the context of cervical cancer screening where unnecessary follow-up can burden health systems. The transition from in-house PCR protocols to validated commercial assays, and more recently the adoption of standardized screening platforms, has been a key driver of improved outcomes.

Experience from the program has also shown that failures in proficiency are sometimes not attributable to the method itself but to how the assay is performed in individual laboratories. This underscores the importance of training, adherence to protocols, and continuous internal quality control, in addition to the external quality assessment provided by the panels. At the same time, some assays have proven exceptionally robust, with laboratories using them almost invariably achieving full proficiency across multiple rounds. Together, these findings highlight that both methodological quality and correct implementation are essential for reliable HPV testing. In this way, the panels have not only measured performance but also shaped the global HPV testing landscape by encouraging uptake of methods that meet internationally agreed standards.

The screening panel was introduced in 2022 to meet the specific needs of cervical cancer screening. Initially, the proficiency cut-offs mirrored those of the typing panel. However, none of the laboratories (n=19) using HC2 were able to detect HPV16/18 at the required threshold of 10 IU/ul, consistent with the assay’s reported detection limit of 100 copies/µl. HC2 is an FDA-approved test, extensively evaluated in randomized controlled trials and used internationally as a comparator assay in HPV test validation.(21) Moreover, Hortlund et al. showed that only 3 of 49 (6%) invasive cancers positive for high oncogenicity HPV types (HPV16/18) were preceded by viral amounts below 100 IU/µl, suggesting that an analytical sensitivity of 100 IU/µl had been sufficient in the original trials.(14) An ongoing international collaborative study where 10 national HPV reference labroatories participate, aims to define the analytical detection thresholds required for an HPV screening test (provisionally 3 IU/ul for HPV16 and HPV18, 25 IU/ul for other oncogenic vaccine-related types, and 100 IU/ul for low oncogenicity types.(unpublished observations) The aim is to use optimal limit of detection thresholds in the screening panel from 2026 onwards. The screening panel would then also serve as an evaluation panel, playing a dual role as both a quality assurance tool for laboratories engaged in cervical screening and an evaluation benchmark against which new HPV assays can be evaluated. An international consensus hearing of experts involved in external assay validation agreed that future assay validations will need to include a component with challenge samples containing defined amounts of virus DNA.(22)

Continued annual proficiency testing remains essential. Looking forward, sustainability will increasingly depend on regional and/or national systems for quality assurance, including proficiency testing. Several countries have already demonstrated that national ownership of quality assurance is feasible and impactful. For example, Peru recently implemented and evaluated a nationwide external quality assessment program for HPV screening, modelled on the internationally standardized program.(23) Similarly, Argentina has for many years distributed the international proficiency panels nationally and to neighboring countries, providing a strong model for regional expansion. These experiences illustrate how the principles of the global program can be adapted and scaled, ensuring that quality assurance reaches laboratories where it is most needed.

By identifying problems early, maintaining international comparability, and building local capacity, the panels can secure the reliability of HPV testing worldwide. As many new assays are developed and screening is expanded in diverse settings, the typing and screening proficiency panels will remain the backbone of a sustainable quality assurance system, to support the goal of cervical cancer elimination.

## Acknowledgments

The success of the HPV proficiency program reflects the combined efforts of many partners. The International HPV Reference Center has coordinated the design, preparation, and evaluation of the panels, ensuring that they are traceable to international standards and accessible to laboratories worldwide. EQUALIS has provided support with distribution of the panels, while the WHO HPV LabNet laid the foundation for the global framework that continues to guide this work. The program has also benefited greatly from the long-standing commitment and expertise of individuals who contributed to the establishment and early development of the HPV LabNet, including Alejandra Picconi, Elisabeth Unger, Suzanne Garland, Iwao Kukimoto, and other members of the network whose sustained engagement was essential in building a robust and internationally trusted quality assurance system. Above all, the program depends on the commitment of the participating laboratories, whose willingness to engage openly in proficiency testing has made it possible to build a shared, international system for quality assurance in HPV testing.

## Author contributions

LSAM: project administration, supervision, validation, writing–original draft. CE: methodology, formal analysis, data curation. EY, CL: methodology. JD: conceptualization, resources, and supervision.

## Data Availability Statement

Results from proficiency panels have all been published as technical reports and most also in peer-reviewed articles (except 2017 and 2024 for genotyping, and 2023 and 2024 for screening which are originally presented here).(5–12) The technical reports are publicly available at https://www.hpvcenter.se/proficiency_panel/.

